# Normative modeling for quantitative brain MRI phenotyping and biomarker discovery for pediatric leukodystrophies

**DOI:** 10.64898/2026.05.22.26353512

**Authors:** Shivaram Karandikar, Anjana Sevagamoorthy, Dabriel Zimmerman, Russell D’Aiello, Lena Dorfschmidt, Katherine Cyr, Benjamin Jung, Elizabeth Levitis, Laura A. Adang, Kaley Arnold, Mariko L. Bennett, Brittany A. Charsar, Carlos A. Dominguez Gonzalez, Francesco Gavazzi, Peter Hong, Jennifer L. Orthmann-Murphy, Sarah T. Pham, Kaleb Kelley, Miriam Lerner, Justine Shults, Nivedita Thakur, Arastoo Vossough, Amy T. Waldman, Aaliyah White, Matthew T. Whitehead, Lisa Emrick, Jamie Fraser, Keith Van Haren, Stephanie Keller, Ali Fatemi, Florian Eichler, Joshua L. Bonkowsky, The Global Leukodystrophy Initiative Clinical Trials Network Workgroup, Jakob Seidlitz, Aaron F. Alexander-Bloch, Adeline Vanderver

## Abstract

**Importance:** Leukodystrophies are a heterogeneous group of genetic disorders affecting the white matter of the brain, often presenting with overlapping clinical features but differing in neuroanatomical involvement. There is a critical need for quantitative tools to characterize disease burden and support diagnosis, severity stratification, and clinical trial readiness.

**Objective:** To characterize shared and distinct neuroanatomical patterns across six genetically confirmed leukodystrophies using anatomical MRI-derived phenotypes benchmarked against brain growth charts, and to assess the utility of this methodological approach for identifying imaging biomarkers of disease severity.

**Design:** Cross-sectional neuroimaging study using retrospective clinical MRI data.

**Setting:** Multicenter study incorporating data from the Global Leukodystrophy Initiative Clinical Trials Network (GLIA-CTN) and control data from the Children’s Hospital of Philadelphia.

**Participants:** The study included 434 MRI scan sessions from 274 patients with genetically confirmed leukodystrophies (Pelizaeus-Merzbacher disease, Metachromatic leukodystrophy, Alexander disease, Aicardi-Goutières syndrome, TUBB4A-related leukodystrophies, and POLR3-related leukodystrophy). Control MRI data (7628 scans from 7205 subjects) were drawn from the Scans with Limited Imaging Pathology cohort at the Children’s Hospital of Philadelphia.

**Exposures:** All MRI scans underwent automated segmentation using deep learning segmentation tools to derive global and regional brain volumes. Normative models of brain development (“brain growth charts”) were generated for the control cohort using generalized additive models for location, scale, and shape. Centile scores were then calculated for leukodystrophy subjects to quantify deviations from typical development.

**Main Outcomes and Measures:** Centile scores for global and regional brain volumes were compared across leukodystrophy subtypes to identify disease-specific neuroanatomical patterns and to evaluate their potential utility for severity stratification.

**Results:** Distinct patterns of neuroanatomical deviation were observed across leukodystrophy subtypes. Certain leukodystrophies showed preferential involvement of specific cortical or subcortical regions, while others displayed more diffuse volume loss. Centile scores demonstrated potential for differentiating disease subtypes and stratifying individuals by severity. Preliminary longitudinal data suggest centile scores may also track progression over time.

**Conclusions and Relevance:** This study demonstrates the feasibility and utility of MRI profiling of individuals with leukodystrophy using anatomical MRI-derived phenotypes benchmarked against brain growth charts. The approach enables data-driven, quantitative characterization of structural brain abnormalities, offering a scalable method for phenotyping, diagnosis, and future use in clinical trials.

**Key Points:** *Question:* In genetically confirmed leukodystrophies, can anatomical MRI measurements benchmarked against brain growth charts identify neuroanatomical patterns that correlate with clinical function and disease severity?

*Findings:* In this cross-sectional neuroimaging study of six leukodystrophies, imaging-derived quantitative phenotypes benchmarked against brain growth charts revealed neuroanatomical patterns of volume loss consistent with previously-reported qualitative changes for each disorder. These patterns of regional volume loss correlated with measures of clinical function, particularly in POLR3-related leukodystrophy, TUBB4A-related leukodystrophy, and Aicardi-Goutières Syndrome.

*Meaning:* Brain growth charts may be a valuable tool for characterizing the patterns of involvement across different leukodystrophies. Furthermore, this approach may facilitate the use of atrophy as a biomarker for assessing disease severity in clinical trials.

## Introduction

Leukodystrophies (LDs) are a heterogeneous group of rare genetic disorders characterized by progressive degeneration of white matter in the central nervous system. Though individually uncommon, collective prevalence of LDs is approximately 1 in 7,500 live births,^1^ a significant contributor to pediatric neurological disease burden. LDs widely in disease mechanism, age of onset, and clinical trajectory. Despite this heterogeneity, many LDs present with overlapping clinical features such as motor dysfunction, cognitive decline, and developmental delay.^2^ However, the extent to which these features converge or diverge across LD subtypes, both clinically and neuroanatomically, remains inadequately understood.

Prior studies have described neuroradiological features of specific LDs, such as frontal predominance in Alexander disease and parieto-occipital involvement in metachromatic leukodystrophy, allowing MRI to support diagnosis through MRI pattern recognition.^3^ Despite this, most prior work is limited to qualitative assessments or small, single-disease cohorts. Comparative analyses across LDs using standardized, quantitative frameworks are lacking.

Recent advances in neuroimaging analysis have introduced robust tools for objective characterization of brain structure from clinical MRIs. SynthSeg, a synthetic data-trained deep learning segmentation algorithm, enables fast and reliable volumetric analysis across diverse imaging protocols and populations.^4,5^ Complementing this, normative brain growth models, or “brain charts”, offer life-spanning reference distributions for brain structure, allowing individual MRIs to be interpreted in terms of age- and sex-adjusted centile scores.^6,7^ These tools open new avenues for benchmarking structural brain variation in rare diseases against normative neurodevelopmental trajectories.

In the present study, we integrate these methods to quantitatively characterize patterns of neuroanatomical deviation in six genetically confirmed LD cohorts. Using centile-based volumetric analysis, our aims are: (1) to identify shared and distinct neuroanatomical signatures across LDs; (2) to test specific anatomical hypotheses, such as the anterior–posterior gradient in Alexander Disease^8^ and the atrophy in Aicardi Goutières Syndrome^9^; and (3) to evaluate the utility of centile scores as potential imaging biomarkers of clinical severity. Together, our findings aim to advance precision phenotyping in leukodystrophies, supporting future efforts in diagnosis, prognosis, and clinical trial design for these challenging disorders.

## Methods

### LD Participants

LD cohorts were identified from the Myelin Disorder Biorepository Project (MDBP) at The Children’s Hospital of Philadelphia (CHOP IRB #14–011236) as part of the Global Leukodystrophy Initiative Clinical Trial Network (GLIA-CTN). The study included subjects with a molecular diagnosis of PMD, confirmed by board certified genetic counselors. Six disorders were selected with brain imaging in at least 20 patients within MDBP: Aicardi-Goutières syndrome (AGS), Alexander Disease (AxD), TUBB4A associated Leukodystrophy (TUBB4A-LD), Metachromatic Leukodystrophy (MLD), Pelizaeus-Merzbacher Disease (PMD), and POLR3-related Leukodystrophy (POLR3-LD).

### Functional Scores in LD subjects

Subjects in were assigned a functional score based on clinical notes from +/- 6 months proximate to clinical MRIs. Subjects with AGS were scored with the AGS Severity Scale, which measures AGS neurologic function and correlates with the Gross Motor Function Measure-88 (GMFM-88).^10^ AxD patients were scored using the Gross Motor Function Classification System (GMFCS). Other LDs were scored using the GMFC-MLD, which has been validated in multiple _LDs._^1^1,1^2^

### Control Participants

Clinical controls were aggregated from brain MRIs obtained CHOP as part of the Scans with Limited Imaging Pathology (SLIP) cohort. SLIP scans were selected through manual review of radiology reports to exclude imaging pathology, as reported previously.^7,13^ Secondary use of clinical scans was determined to be exempt from review by CHOP IRB because it repurposes preexisting and deidentified clinical data.^14^

### Image Processing

All scans were converted from DICOM to NIfTI using HeuDiConv^15^ and curated into Brain Imaging Data Structure (BIDS) format using CuBIDS^16,17^ (https://github.com/BGDlab/chop-bgd-image-curation). Cortical surface analysis and segmentation were performed using *recon-all-clinical* in FreeSurfer v7.4.1.^4,5,18,19^ Measures of global and regional volumes were output for both cortical and subcortical structures (**eFigure 1**). Quality control of processed images used the Euler characteristic and automated QC measures output by SynthSeg+.^5^ Scans were excluded from further analysis if mean Euler characteristic <-60 or the SynthSeg+ QC scores <0.65.

**Figure 1:**
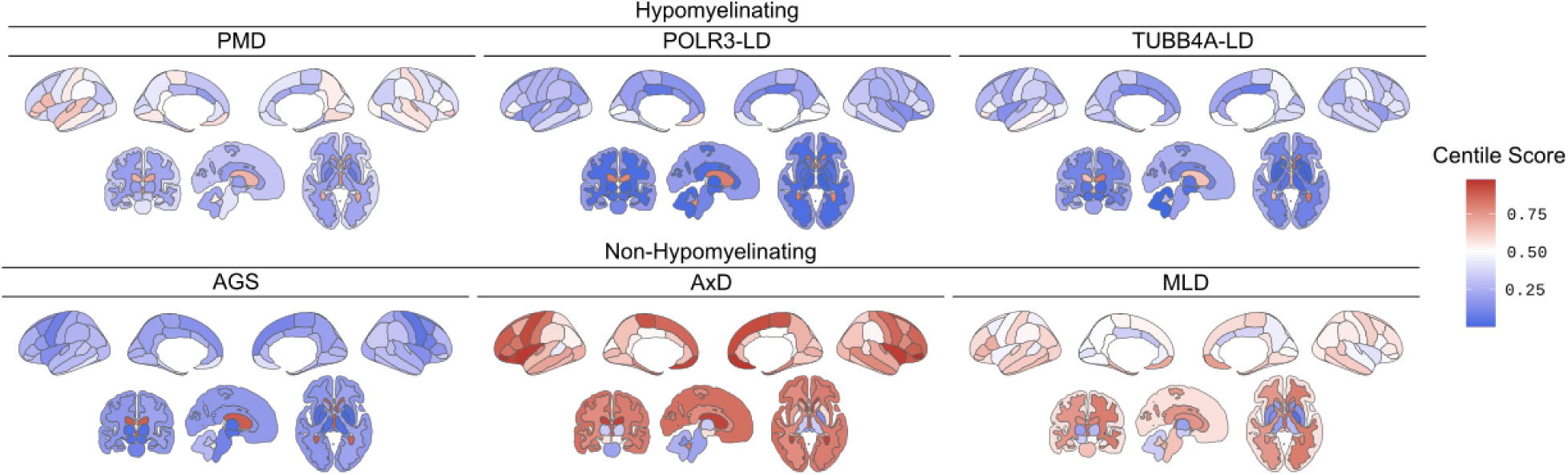
Anatomical MRI profiles for six leukodystrophies benchmarked against brain growth charts. Median centile scores for regional cortical volumes (top panels) and global/subcortical volumes (bottom panels) by LD subtype. Brain maps display the distribution of regional brain volume centiles across LD subtypes, differentiating between higher (red) and lower (blue) median centiles. Regional cortical volumes were derived for 34 regions per hemisphere from the gyrus-based Desikan-Killiany parcellation. Subcortical volumes were derived from the aseg segmentation. Global volumes include total gray and white matter per cerebral hemisphere. See **e**Figure 1 for image key. Abbreviations: AGS, Aicardi-Goutières syndrome; AxD, Alexander Disease; MLD, Metachromatic Leukodystrophy; PMD, Pelizaeus-Merzbacher Disease; POLR3-LD, POLR3-related Leukodystrophy; TUBB4A-LD, TUBB4A-related Leukodystrophy

### Normative Modeling

Growth charts for clinical controls were generated using generalized additive models for location, scale, and shape (GAMLSS).^20^ Imaging phenotypes were fitted to the generalized gamma distribution with age and sex as fixed effects, and scanner ID as a random effect, as previously described.^6^ Normative trajectories were plotted to display percentile curves by sex over the age range. To produce a single observation per scan session, the median volume of each imaging phenotype was calculated across all available sequences as previously described.^5,21^ This approach limited the influence of outliers resulting from sequence-specific artifacts and errors in processing.

Centile scores for the LD cohort were predicted using growth chart models derived from clinical control data. Since a majority of LD scan sessions originated from external scanner sites not included in the SLIP cohort, site effects were estimated by generating predictions for each LD scan session across all scanners represented in the model’s random effects. The median centile score across scanner sites was then calculated for each session to produce a single centile estimate per observation.

### Correlation of centiles with functional scores

We applied linear mixed-effects modeling to assess how centile-based deviations in regional and global brain volumes related to disease severity as defined by functional scores, while accounting for age, sex, and other covariates. Prior to modeling, centile scores were transformed to a standard normal scale using the probit transformation. A random effect at the participant level was included to account for repeated observations across scan sessions.

Statistical significance was determined using false discovery rate (FDR) correction (p.adj < 0.05).

## Results

### Cohort characteristics

Data from MDBP consisted of 838 scan sessions from 372 subjects. Following processing and QC, the LD cohort consisted of 434 scan sessions from 274 subjects (mean age=7.18 years). The SLIP cohort used to generate growth charts consisted of 7628 scan sessions from 7205 subjects (mean age=11.21 years). See **Table 1**.

**Table 1:**
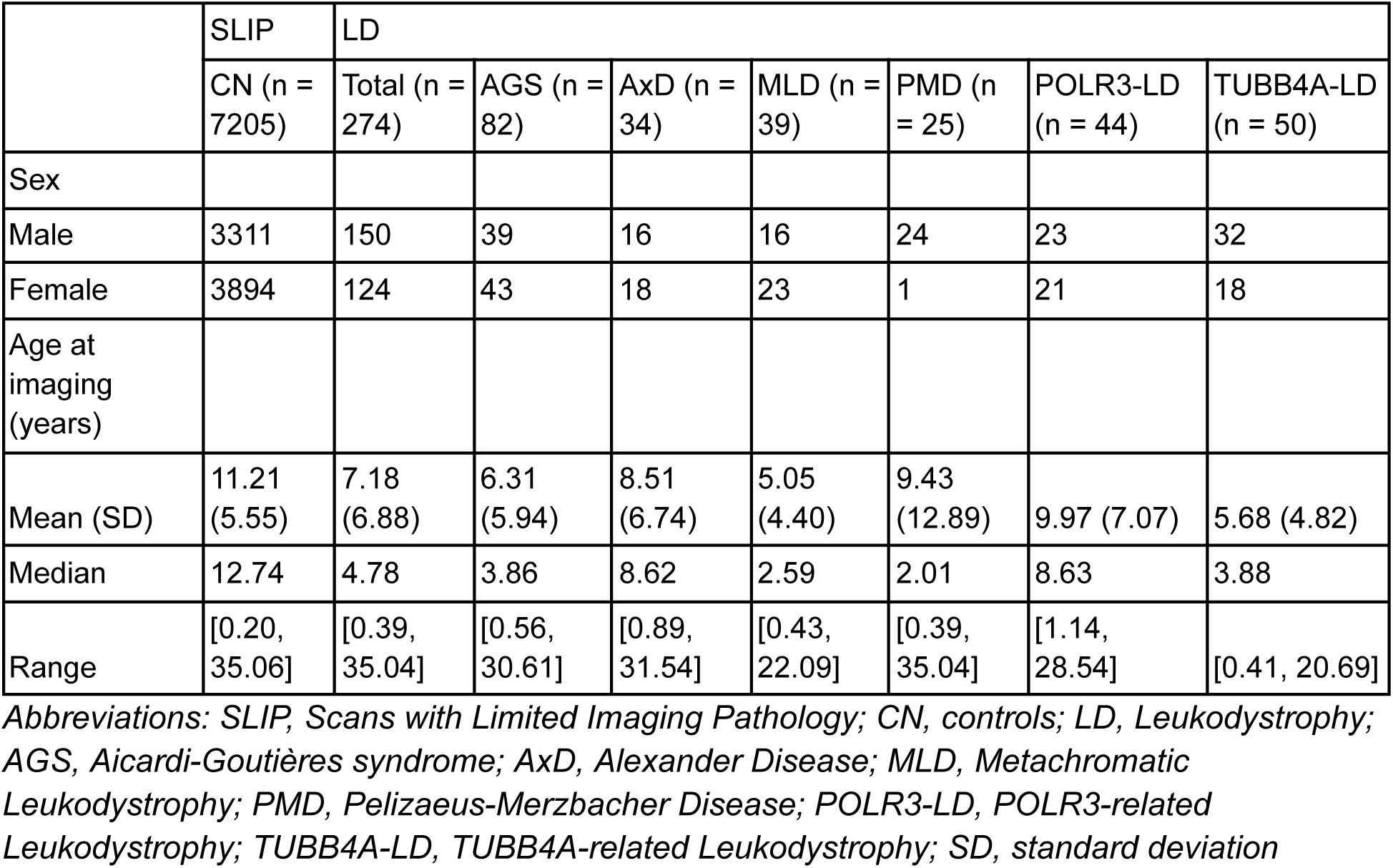
Demographic information in study cohort.

### Global and subcortical volume effects

Quantitative centile-based volumetric analysis revealed distinct neuroanatomical deviation patterns across the six LD subtypes. AGS, TUBB4A-LD, PMD, and POLR3-LD exhibited widespread volume loss, with most cortical and subcortical regions scoring below the 25th centile relative to age- and sex-matched controls. In contrast, AxD and MLD demonstrated relatively increased volume in many regions, with median centile scores frequently above the 50th percentile. Notably, all six subtypes exhibit relatively increased ventricle volume, consistent with global atrophy and tissue reorganization as a result of pathology. The volumes of subcortical structures such as thalamus and basal ganglia were markedly decreased in AGS, PMD, and TUBB4A-LD. These centile profiles support both convergent and divergent anatomical involvement across leukodystrophies and highlight the utility of normative centile scoring for differentiating subtypes and assessing regional vulnerability (**Figure 1, eFigures 2-5**).

**Figure 2:**
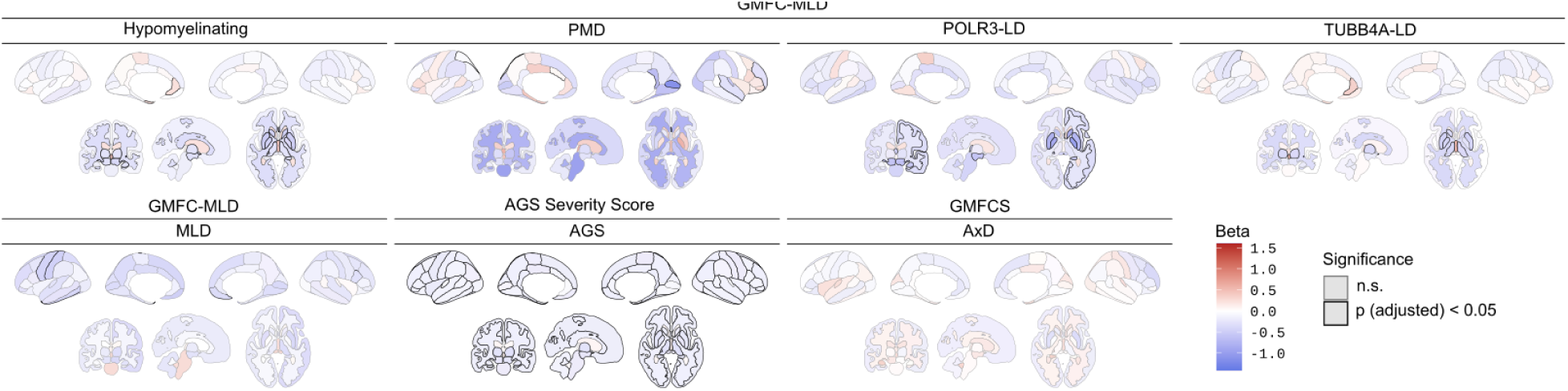
Neuroanatomical correlates of gross motor function in leukodystrophies. Brain maps of fixed effect estimates from linear mixed effects models relating clinical measures of gross motor function—GMFC-MLD, GMFCS, or AGS Severity Score—to regional and global brain volume centiles. For GMFC-MLD, analyses were performed on all hypomyelinating leukodystrophies, and separately for TUBB4A-LD, PMD, MLD, and POLR3-LD groups. Analyses using GMFCS and the AGS Severity Scale were conducted for AxD and AGS respectively. Standardized beta coefficients are visualized for each region, reflecting the association between measures of motor function and brain volume, with regions outlined in bold indicating statistical significance after FDR correction. See **e**Figure 1 for image key. Abbreviations: GMFC-MLD, Gross Motor Function Classification in MLD; GMFCS, Gross Motor Function Classification System; AGS, Aicardi-Goutières syndrome; AxD, Alexander Disease; MLD, Metachromatic Leukodystrophy; PMD, Pelizaeus-Merzbacher Disease; POLR3-LD, POLR3-related Leukodystrophy; TUBB4A-LD, TUBB4A-related Leukodystrophy

### Cortical volume effects

Regional cortical centile maps revealed marked differences in cortical volume preservation and atrophy across LD subtypes. AGS, TUBB4A-LD, PMD, and POLR3-LD demonstrated globally reduced cortical volumes across regions, with most regions falling below the 25th centile. These findings are consistent with diffuse cortical involvement and neurodevelopmental impairment common to these disorders.^3^ In contrast, AxD exhibited a striking anterior-predominant pattern of relative volume preservation, with many frontal and temporal regions exceeding the 75th centile. This aligns with known frontal predominance of AxD pathology.^3,8^

MLD showed widespread but milder reductions in cortical volume, with most regions between 25th and 50th centiles, suggesting more uniform cortical impact. While overall volume loss was less pronounced in MLD compared to other subtypes like PMD or AGS, several regions (including parietal and temporal cortices) showed consistent deviations from reference brain charts. Together, these cortical profiles further support disease-specific anatomical signatures and highlight the sensitivity of centile-based analysis in capturing nuanced regional vulnerability.

### Regional Brain Associations with Functional Scores by Diagnosis

To systematically map neuroanatomical correlates of motor disability in pediatric leukodystrophies, we identified associations between GMFC or similar functional scores and regional brain volumes across both pooled and diagnosis-specific cohorts (**Figure 2, eTables 1-7**). GMFC provides a clinically validated, cross-disease metric of motor impairment severity, making it a powerful tool for identifying shared and divergent anatomical substrates of motor dysfunction.

#### Hypomyelinating Leukodystrophies

Among individuals with hypomyelinating disorders (including PMD, POLR3-LD, and TUBB4A-LD), GMFC scores were associated with 27 regional volumes. Strongest effects were found in right putamen (β = –0.49, p.adj = 4.6×10^-4^), left putamen (β = –0.47, p.adj = 2.2×10^-4^), right accumbens (β = –0.44, p.adj = 6.3×10^-3^), right caudate (β = –0.44, p.adj = 2.5×10^-4^), and left caudate (β = –0.43, p.adj = 3.2×10^-4^) (**Figure 2, eTable 1**). These data emphasize a shared subcortical burden among myelin-disruptive conditions affecting motor function.

#### POLR3-Related Leukodystrophy (POLR3-LD)

In POLR3, GMFC severity was significantly associated with 16 regional volumes, primarily in subcortical and limbic structures. Notable reductions were observed in left pallidum (β = –0.97, p.adj = 6.2×10^-3^), right pallidum (β = –0.84, p.adj = 2.8×10^-2^), left caudate (β = –0.79, p.adj = 1.7×10^-2^), right putamen (β = –0.77, p.adj = 1.2×10^-2^), and left amygdala (β = –0.74, p.adj = 7.3×10^-3^) (**Figure 2, eTable 2**), underscoring POLR3-LD’s mixed extrapyramidal and cognitive presentation.^22,23^

#### TUBB4A-Related Leukodystrophies (TUBB4A-LD)

In TUBB4A, GMFC severity was significantly associated with 16 regional volumes. Prominent findings included third ventricle enlargement (β = +0.53, p.adj = 9.2×10^-3^), reduced volume right accumbens (β = –0.48, p.adj = 3.2×10^-2^), right thalamus (β = –0.38, p.adj = 3.2×10^-2^), right putamen (β = –0.41, p.adj = 3.5×10^-2^), and global cerebral white matter (β = –0.39, p.adj = 4.1×10^-2^) (**Figure 2, eTable 3**), reflecting characteristic subcortical and white matter degeneration.^24^

#### Pelizaeus-Merzbacher Disease (PMD)

In PMD, GMFC severity was associated with 7 cortical regions. Significant effects included reduced volume in right pericalcarine cortex (β = –1.13, p.adj = 4.5×10^-29^), right cingulate isthmus (β = –0.65, p.adj = 5.3×10^-27^), right superior temporal gyrus (β = –0.29, p.adj = 4.9×10^-22^), right rostral middle frontal gyrus (β = –0.29, p.adj = 1.2×10^-21^), and right lateral orbitofrontal cortex (β = +0.08, p.adj = 3.9×10^-21^) (**Figure 2, eTable 4**). These associations highlight distinct cortical correlates of motor severity in PMD, particularly in visual, frontal, and limbic regions.

#### Metachromatic Leukodystrophy (MLD)

In MLD, GMFC severity was significantly associated with 6 cortical phenotypes: left temporal pole (β = –0.50, p.adj = 2.7×10^-2^), left postcentral gyrus (β = –0.44, p.adj = 2.4×10^-2^), left precentral gyrus (β = –0.43, p.adj = 4.5×10^-2^), left inferior temporal gyrus (β = –0.41, p.adj = 2.2×10^-2^), right medial orbitofrontal cortex (β = –0.35, p.adj = 4.8×10^-2^), and right pars opercularis (β = –0.29, p.adj = 1.1×10^-2^) (**Figure 2, eTable 5**). These effects localize to posterior and sensorimotor cortex, consistent with the clinical profile of MLD.^24^

#### Alexander Disease (AxD)

In AxD, GMFCS severity was not associated with any single region after correction for multiple comparison. The strongest association was found in the left putamen (β = –0.33, p.adj = 5.0×10⁻²) (**Figure 2, eTable 6**).

### Regional Brain Associations with AGS Scores in AGS Patients

To complement the transdiagnostic GMFC analysis, we examined brain-behavior relationships specific to Aicardi-Goutières Syndrome (AGS) by modeling associations between total AGS Severity Scores and regional brain volumes. Unlike GMFC, which primarily indexes motor disability, AGS scores captures a broader spectrum of clinical impairment (including cognitive, sensorimotor, and systemic features) characteristic of AGS.^9^ Because the original AGS scale is reversed compared to GMFC (i.e., lower scores indicate greater disease severity), AGS scores were rescaled for consistency with GMFC analyses.

Linear mixed-effects modeling identified 108 brain phenotypes significantly associated with AGS severity after FDR correction (p.adj < 0.05) (**Figure 2, eTable 7**). Strongest associations were observed in subcortical and diencephalic structures. Higher AGS severity was also associated with lower subcortical centile scores (**Figure 2, eTable 7**). Cortical associations with AGS severity were also significant, particularly in limbic and paralimbic regions such as left posterior cingulate cortex (β = −0.17, p.adj = 2.0×10^-6^), left insula (β = −0.18, p.adj = 2.2×10^-6^), and right isthmus cingulate (β = −0.16, p.adj = 4.2×10^-6^) (**Figure 2, eTable 7**). These effects converge on brain systems involved in motor coordination, sensory integration, limbic regulation, and deep gray matter relay, consistent with the clinical phenotype of AGS.

Together, these findings confirm that greater AGS clinical severity is tightly linked to anatomical disruption across both cortical and subcortical systems, with robust effects in thalamic, basal ganglia, and diencephalic structures. The alignment of AGS-specific severity scores with regional volume loss in clinically relevant areas reinforces the construct validity of this diagnostic instrument. Moreover, these neuroanatomical correlates support the utility of the AGS Severity Score for patient stratification, progression monitoring, and treatment response evaluation in natural history studies and early-phase interventional trials targeting pediatric neuroinflammatory encephalopathies.

## Discussion

Normative modeling applied to routine clinical MRIs can provide a standardized framework for characterizing neuroanatomical variation across pediatric LDs. By benchmarking individual brain volumes against age- and sex-adjusted reference distributions, centile-based measures identified both shared and syndrome-specific patterns of structural deviation and showed associations with clinical measures of disease severity. These findings suggest that quantitative deviation from typical brain development captures clinically meaningful aspects of disease burden that are not accessible through conventional qualitative imaging assessments.

Expert analysis of imaging patterns has long been recognized as an important diagnostic tool in LDs.^3^ Despite the success of pattern recognition in directing diagnostic testing for LDs,^25,26^ this approach is insufficiently quantitative to permit imaging to be used for stratification or as an outcome biomarker for candidate drug trials. Use of automated imaging approaches holds potential to overcome previous methodological hurdles.

In this study, we applied automated approaches to quantify volume loss relative to age matched controls in a series of LDs. Importantly, findings aligned with expected volume changes in specific disorders. For example, our findings replicate known atrophy across hypomyelinating LDs, in particular for POLR3-LD and TUBB4A-LD, greatest in striatal gray and cerebellum.^22,27–29^ Similarly, AGS showed diffuse atrophy of supra and infratentorial structures with corresponding ventriculomegaly.^9^ Conversely, Alexander disease showed supratentorial megalencephaly and infratentorial atrophy, consistent with clinical impressions.^8^ MLD, a lysosomal storage disorder, evidenced moderate increase in volume of cerebral white matter.

Critically, the degree of atrophy correlated with clinical function in several disorders. For hypomyelinating disorders, in particular POLR3-LD and TUBB4A-LD, degree of atrophy of deep gray nuclei corresponded to worsening clinical function as measured by GFMC-MLD. Notably, this was not true in PMD. In AGS, diffuse atrophy was prominently associated with functional outcomes measured by the AGS Severity Scale. Volumetric changes did not notably correlate with outcomes in AxD and MLD.

Of note, the pattern of atrophy and its association of clinical function appeared to be lateralized in several analyses. For instance, only lower volumes in the left hemisphere primary motor cortex were significantly correlated with lower motor function in MLD. While suggestive, these findings may reflect a combination of biological variability and methodological factors, and should therefore be interpreted cautiously pending validation in larger and longitudinal datasets.

An important area of innovation of the present study is the repurposing of clinically-acquired brain MRIs for automated image analysis. Given the difficulty and expense of prospective neuroimaging studies, repurposing clinical MRIs has potential to dramatically increase data available for biomarker discovery. Nonetheless, LD sample size remained a constraint, not only limiting statistical power in PMD but also preventing investigation of gene-specific patterns of involvement in genetically heterogeneous LDs such as MLD, AGS, and POLR3-LD.^30^ Increased participation in multi-site imaging consortia will lead to increased sample sizes and statistical power to test such hypotheses. The incorporation of “clinical controls” from all LD consortium sites in future studies would improve statistical harmonization to brain growth charts and could be accomplished at-scale with AI-assisted review of neuroradiology reports.^13,31^

There are several limitations to consider in the interpretation of the present study. While repurposing clinical MRIs holds great potential, specific imaging markers should still be validated in controlled research settings to minimize ascertainment bias, sequence- and scanner-related heterogeneity prior to being deployed in clinical trials. Furthermore, lack of extensive longitudinal data limited our ability to characterize within-subject disease progression. Future longitudinal studies could clarify relationships between structural and functional change over time. Finally, the present findings are bound by the age range of the current sample. Certain neuroanatomical deviations may not be captured in our predominantly pediatric cohort.

Despite these limitations, the present study establishes the feasibility of automated imaging markers in this cohort of rare pediatric LDs. The data additionally suggest that for a subset of LDs, measures of atrophy correspond to clinical severity and may be useful as a stratification biomarker. Future work should focus on prospective validation and longitudinal analyses to determine the sensitivity of these measures to disease progression and therapeutic effects. If further validated, this approach may help advance precision phenotyping and biomarker-driven trial design in pediatric neurology.

## Disclosures

SKa, KC, BJ, EL, DZ, and AA-B have an inventorship interest in intellectual property licensed by the Children’s Hospital of Philadelphia to Centile Bioscience. JSe and AA-B hold shares in and JSe is a director of Centile Bioscience. LAA is an advisor and site sub-investigator for Takeda Pharmaceuticals and Orchard Therapeutics. FG receives research support from Ionis Pharmaceuticals. RD has received research support from Takeda Pharmaceuticals. AVo is a research consultant to Syneos Health and advisor to DeepSight. JLB has clinical trials with Calico and Ionis Pharmaceuticals; consulting with Calico and Ionis Pharmaceuticals; writing content for UpToDate; stock in Orchard Therapeutics; and royalties from BioFire and Manson Publishing. JO-M has grant support from NIH and NMSS; consultant with Ionis Pharmaceuticals and Savanna Bio; site PI for Vigil Neuroscience. KK has received compensation from Biogen Inc, Syneos Health, and IQVIA. SKe has received clinical trial support from Ionis Pharmaceuticals.

## Supporting information

eFigure 3

eFigure 4

eFigure 5

eFigure 2

eFigure 1

eTables

## Data Availability

The participant-level data cannot be made publicly available as they contain protected health information from pediatric patients.

## Acknowledgements

This work was funded by NINDS U01NS106845, NIMH R01MH134896, NIMH R01MH133843, NINDS/NCATS U54NS115052, NINDS R61NS135584 and the CHOP Research Institute.

## Notes

### Author Declarations

The Institutional Review Board of the Children's Hospital of Philadelphia gave ethical approval for the usage of data collected from the Myelin Disease Biorepository Project (IRB 14-011236), and waived ethical approval for the usage of clinical brain MRIs as part of the reference cohort (IRB 20-017870).

