## Supplementary figures and images for "Normative modeling for quantitative brain MRI phenotyping and biomarker discovery for pediatric leukodystrophies"

### eFigure 1

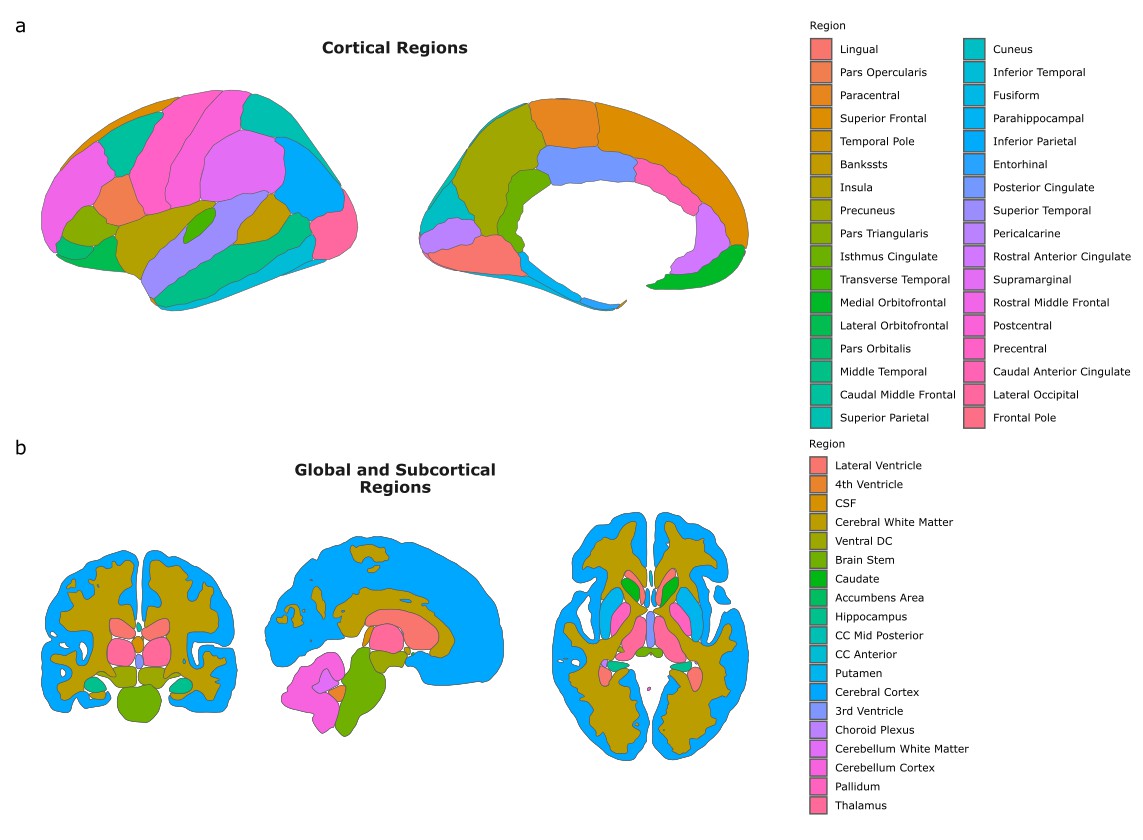

### eFigure 2

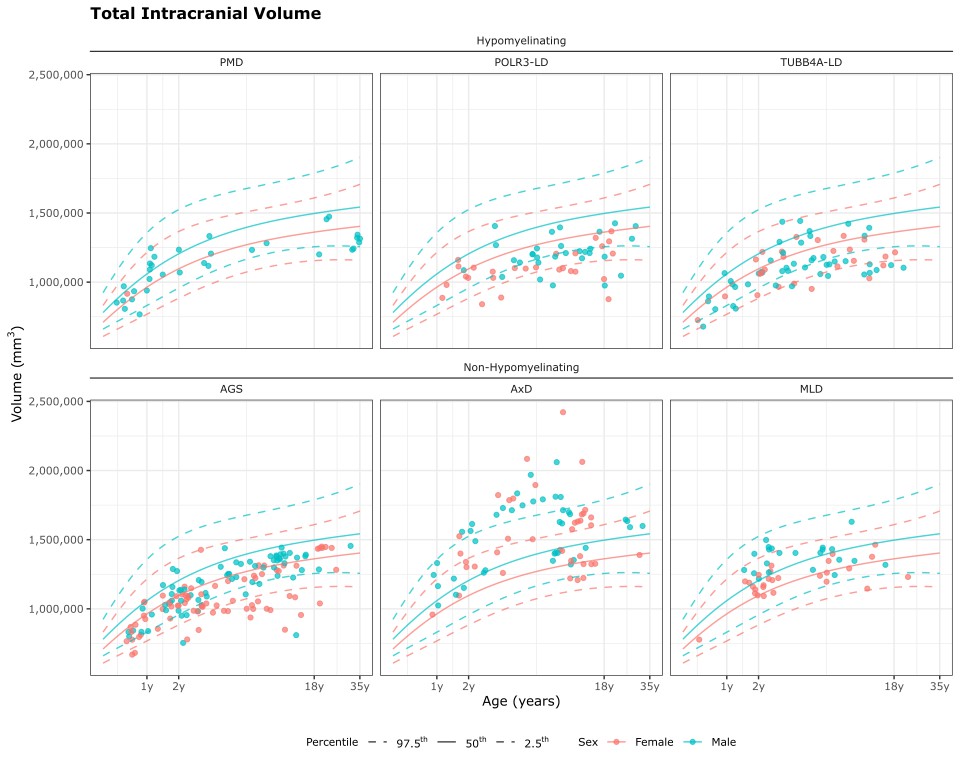

### eFigure 3

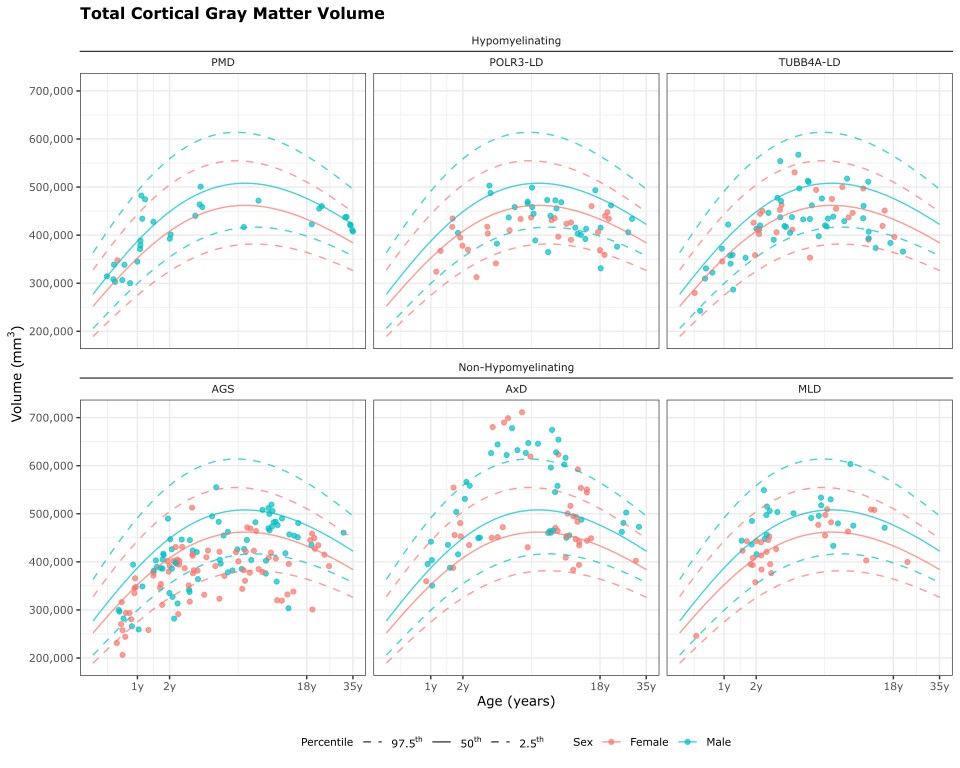

### eFigure 4

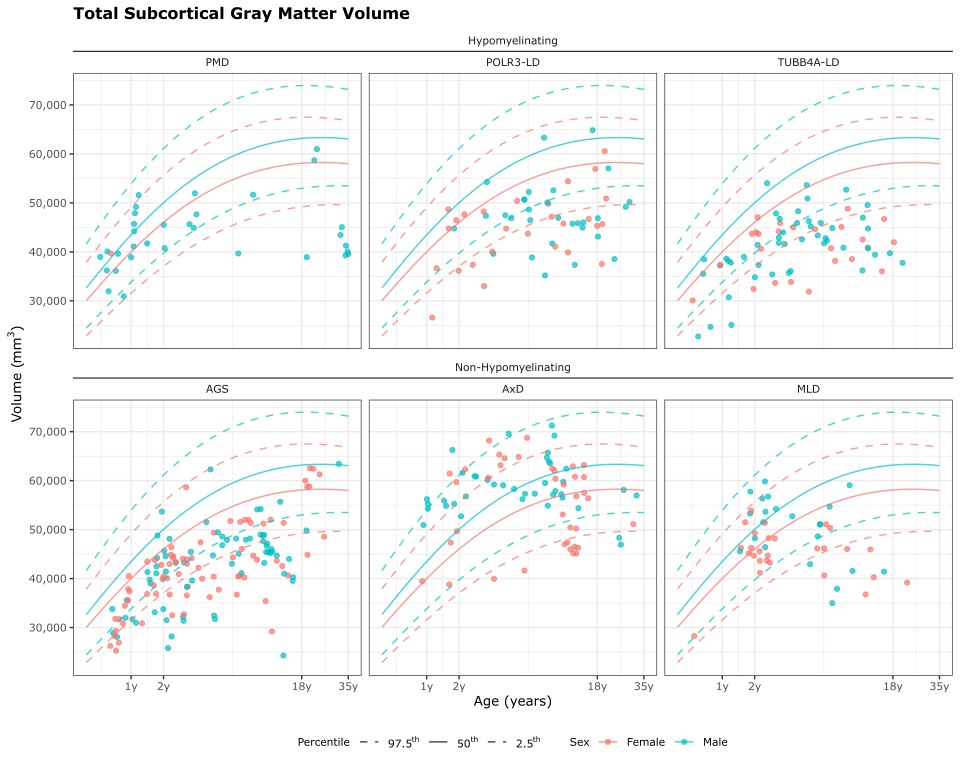

### eFigure 5

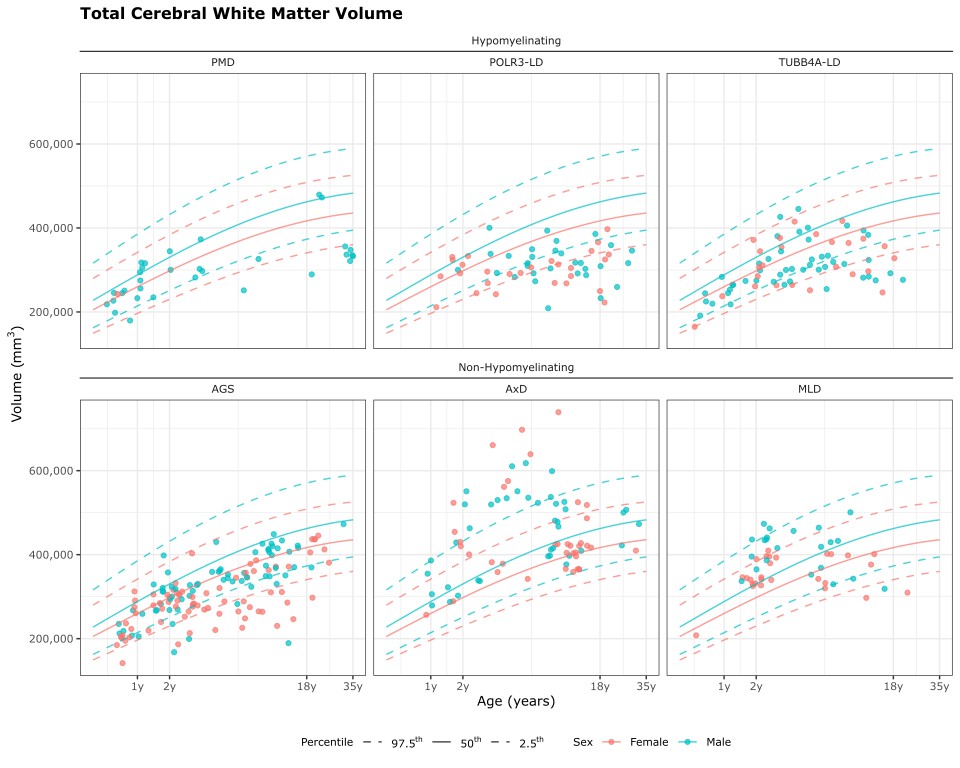
